# Supporting carers: Study protocol of a meta-review of psychosocial interventions for carers of people with cancer

**DOI:** 10.1101/2024.01.30.24302029

**Authors:** Bróna Nic Giolla Easpaig, Bronwyn Newman, Judith Johnson, Rebekah Laidsaar-Powell, Ursula M Sansom-Daly, Lucy Jones, Lukas Hofstätter, Eden G Robertson, Stephen Mears, Kabir Sattarshetty, Reema Harrison

## Abstract

**Background:** While there is a clear need for psychosocial interventions that promote cancer carer wellbeing, the corresponding evidence base is disparate, complex and difficult for end-users to navigate and interpret. Carers remain under-supported with a lack of dedicated, effective, evidence-based programs. We will conduct a meta-review to synthesise this evidence and determine the state of science in this field.

**Objectives:** This study aims to address the question of: “What psychosocial interventions are available to promote the wellbeing of carers for people with cancer?”

**Methods:** A meta-review will synthesise relevant reviews of psychosocial interventions that have been developed and/ or evaluated with carers for people with cancer. Four electronic databases (PsychInfo, Medline, CINAHL and Cochrane Database of Systematic Reviews) will be searched for reviews published between Jan 2013 and Dec 2023. A team-based approach will be taken to screening and assessment of the returned records against the eligibility criteria to determine inclusion. Included reviews will be critically appraised using JBI Critical Appraisal Checklist for Systematic Reviews and Research Syntheses. Relevant data of study characteristics, carer and patient populations, intervention details and psychosocial outcomes will be extracted, synthesised and the findings will be presented in a narrative format. This study is registered with the International Prospective Register of Systematic Reviews (reference: CRD42023403219).

**Results:** It is anticipated that the study will be completed by April 2024.

**Conclusion:** Ensuring that carers have access to evidence-based programs which promote their wellbeing as they care for loved ones is critical. This meta-review will contribute to program development and translation efforts through providing a clear picture of the cancer carer intervention evidence-base, identifying notable strengths, weaknesses, and gaps across the literature. The findings are anticipated to offer future directions to advance research in the field.

## Introduction

Family and friend carers serve as a core, yet under acknowledged members of the health team in coordinating and providing care for loved ones diagnosed with cancer [1]. The partners, parents, siblings, children and friends of patients often fulfil this role; a role which spans pragmatic, clinical and emotional domains of support [2-4]. Shifts in oncology care delivery, increasingly towards outpatient, community and home settings have widened the scope of carers’ roles and responsibilities [1, 4-6]. Carers of people with cancer may assume significant responsibilities in not only coordinating and organising care, but in providing direct clinical care too (e.g., administering medications) [4, 6].

Becoming a carer is a role that many feel unprepared for and are overwhelmed by, with implications for health and wellbeing [7, 8]. Carers experience depression, anxiety and distress, commonly at higher rates than the general population [9-11]. Further, both the quality of care and clinical outcomes of the patient are linked to the wellbeing of carers [6]. There has been a growing interest in identifying psychosocial interventions that may be effective in supporting this population [7, 12].

While there appears to be a large volume of literature reporting on studies of psychosocial interventions for cancer carers, this body of work is complex, fragmented, and it is challenging to draw clear conclusions about the evidence for specific types of programs or carer groups. The result is that carers remain under-supported, with limited evidence of the effective interventional approaches. Key issues within the cancer carer evidence-base that limit advancement in practice are the divergent scope and focus of current interventions (e.g. in person therapy for carers, web-based interventions for patient-spouse dyads [13], compared with psychological interventions for parents of children and adolescents with chronic illness [14]) all of which may be included in a single review.

A potential weakness in the current literature is that interventions have not necessarily been designed for carers as the target primary population; programs may be developed for patients and extended to include carers [15]. In such cases, carer-specific needs may not be met. Additionally, the nature of relationships between carers and the person being cared for are not always sufficiently considered in the collation of intervention evidence. While findings suggest that distress and stress can arise while providing health care and with a loved one being ill [3, 16], this appears under-explored. These limitations alongside the divergences in the literature described pose barriers for end-users of this evidence [17].

### Review aim and question

Undertaking a meta-review was identified as a useful first step in addressing the abovementioned barriers to enable the development of targeted interventions that may offer greater impacts to reduce carer distress and enhance support [18]. A meta-review offers a means to develop an overall, coherent picture of a large volume of evidence [19, 20], that is useful for those wishing to navigate this literature and identify the evidence relevant to them. The aim of this study is to synthesise the evidence from reviews of psychosocial interventions designed to support the wellbeing of carers of people with cancer.

This study will address the question of:

What psychosocial interventions are available to promote the wellbeing of carers for people with cancer, as reported in the evidence from reviews?

## Methods

Meta-review was employed as a method that offers a systematic and rigorous approach to the identification and review of relevant evidence in the form of various types of reviews [21]. The protocol for this study is registered with the International Prospective Register of Systematic Reviews (reference: CRD42023403219). In the absence of a method-specific protocol reporting framework, the Preferred Reporting Items for Systematic Reviews and Meta-Analysis Protocols (PRISMA-P) 2015 guidelines are used to report this study protocol (please see Supplementary file 1)[22].

### Eligibility criteria

The PICO framework was used as a basis for developing the eligibility criteria. The following criteria will be used to determine study inclusion.

### Information sources

Systematic searching will be undertaken of the PsychInfo, Medline, CINAHL and Cochrane Database of Systematic Reviews databases. Additionally, the reference lists of relevant reviews will be audited to identify other potentially eligible reviews. The search will cover a ten-year period from the 1^st^ of January, 2013 until the 1^st^ of December 2023.

### Search strategy

The search strategy for the databases listed was developed via consultation with a medical research librarian. The search was developed based on the search strategy employed in the Treanor et al.’s (2018) Cochrane review of the psychosocial interventions for informal carers of people living with cancer [23], and informed by concepts encompassed in Fletcher et al. (2012) model of the cancer family carer experience [24]. Search terms updated as required, including for terms related to study type (e.g. “systematic review”). The search period was selected to identify recently published reviews and capture the current evidence landscape. The finalised search strategy uses a combination of Medical Subject Heading terms and key words and as an example, the strategy developed for the Medline (Ovid) database is included in Supplementary file 2.

### Selection process

Records retrieved from the searches will be imported into EndNote X9 (citation management software) [25], and duplicates subsequently removed. The remaining records will then be uploaded to Covidence (literature review management tool) [26], which will be used to manage the screening of records. The titles, abstracts and keywords of records will be screened by one of the team members against the criteria to determine inclusion. The full texts for included records will then be retrieved and two team members will independently assess each text against the eligibility criteria. Disagreements will be resolved by team discussion, with discrepancies discussed with a third team member until resolution is reached. The search results will be documented and reported using the Preferred Reporting Items for Systematic Reviews and Meta-Analyses (PRISMA) guidance [27], with adaptations made as needed to reflect the meta-review method.

### Data collection process

A draft data extraction template will be developed, and two team members will independently extract data for a shared 10% of the included reviews to identify any amendments needed to the template. Once finalised, a team-based approach to data extraction will be taken whereby data will be extracted by one team member, all of which will be subsequently cross-checked by another member of the team. Discrepancies will be resolved via discussion. Microsoft Excel will be used to manage the data extraction process [28]. Data will be extracted in the areas of: study characteristics, populations of carers and patients, intervention details, outcomes of interest. See Table 2 for full details.

**Table 1.**
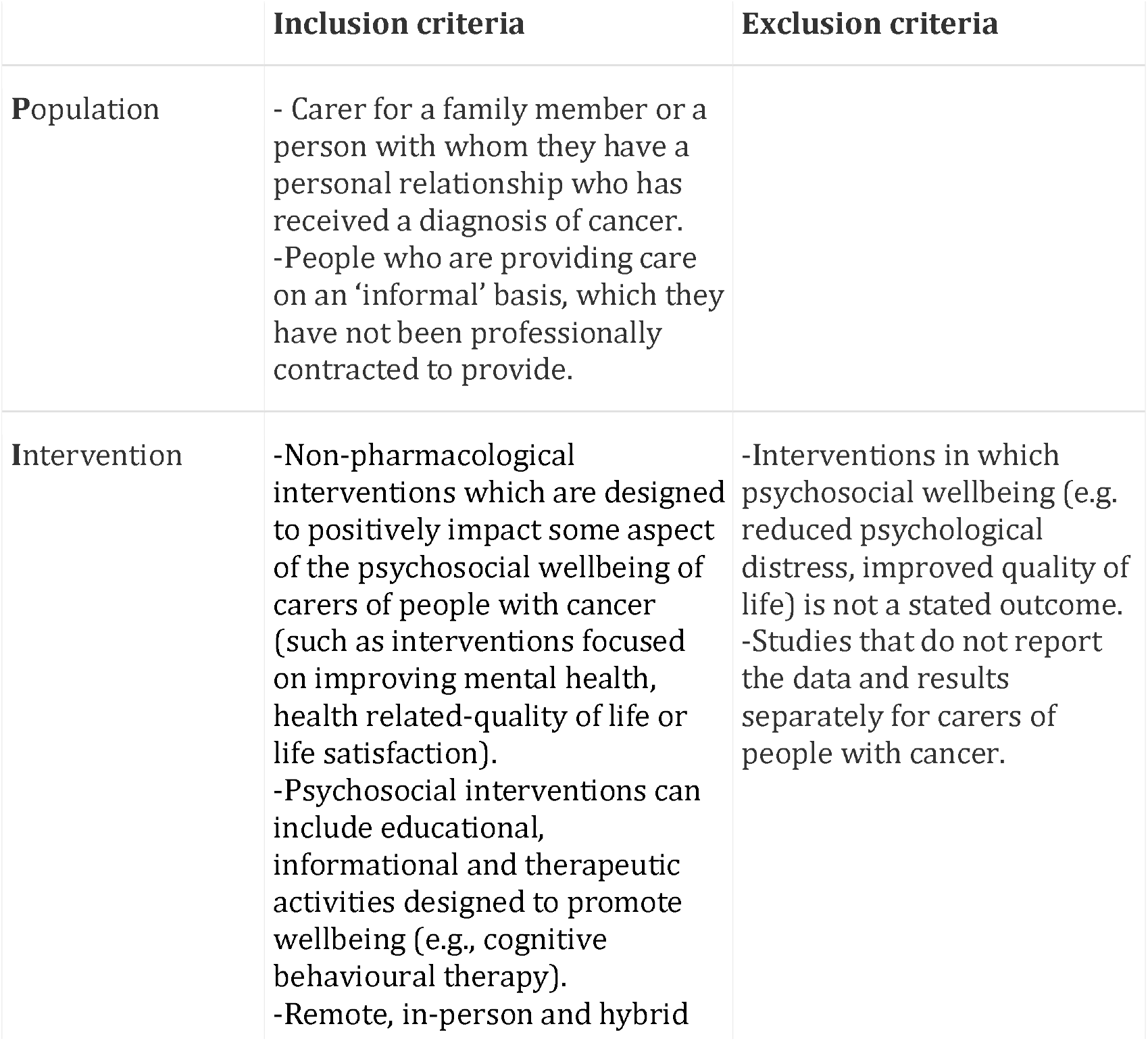
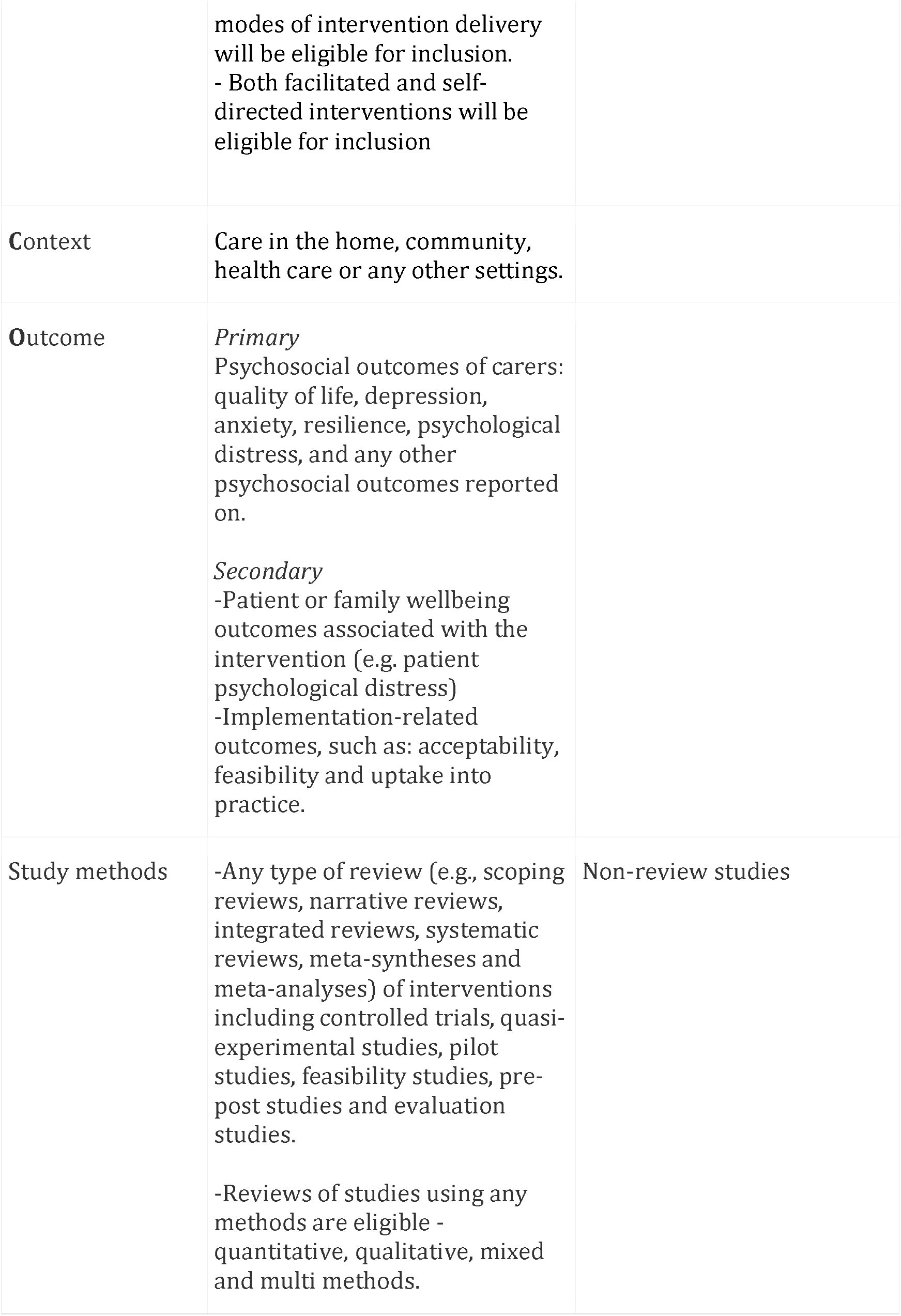
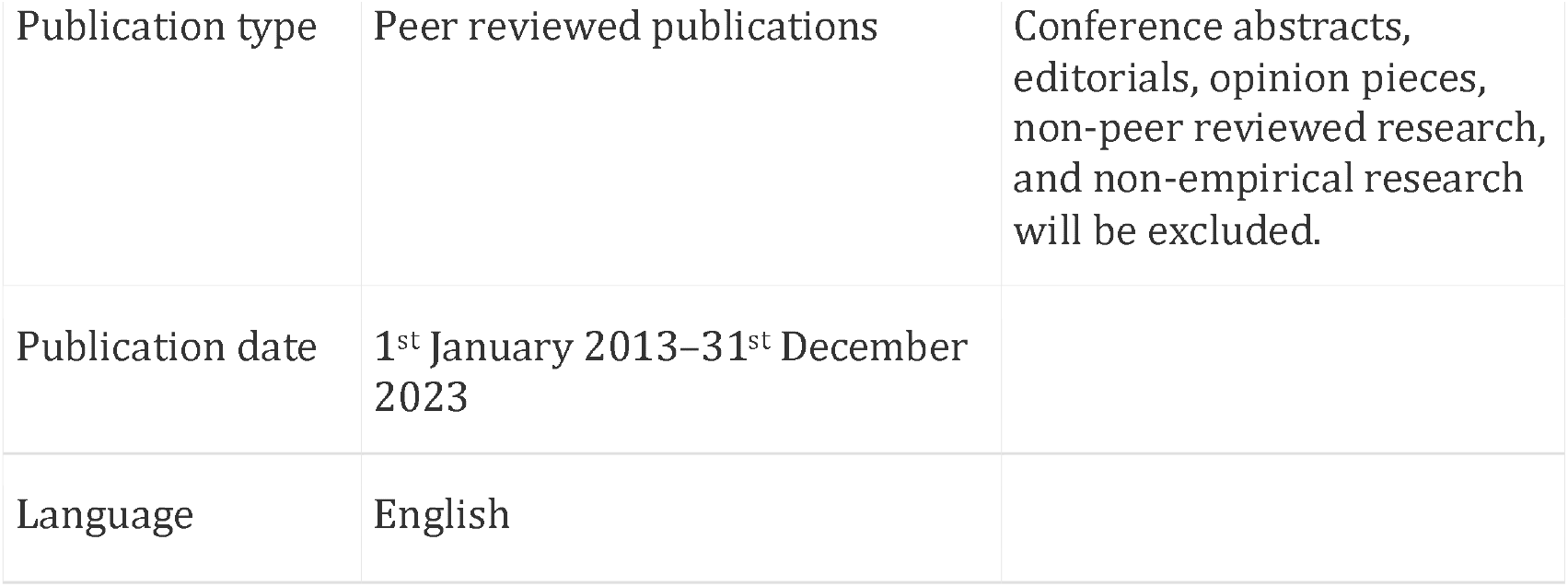
Eligibility criteria.

**Table 2.** Data items for extraction.

| Areas of data collection | Data item details |
| --- | --- |
| **Study characteristics** | Year, review aim(s), types of review, any geographical restrictions of review, condition(s) of those cared for methods, study types/ designs included in review, total number of included studies (and articles, if different), participants total number/ sample size details, search period, synthesis method, critical/ quality appraisal tool used and any other notable details |
| **Population** | Population focused on in the review, target carer population, target, patient population and any other notable details. |
| **Intervention details** | Details about of intervention development, mode of delivery, theoretical bases, settings, facilitators, details about frequency/duration/length of interventions, and any other notable details. |
| **Outcomes of interest** | Carer psychosocial outcomes: quality of life, depression, anxiety, resilience, psychological distress, and any other psychosocial outcomes reported on.<br>Implementation-related outcomes: acceptability, feasibility and uptake into practice and other implementation-relevant outcomes.<br>Patient or family wellbeing outcomes associated with the intervention such as patient psychological distress. |

### Data synthesis

The extracted data will be collated and organised. A narrative approach will be taken to describe the results, study characteristics, populations, interventions, outcomes and any other details of interest. Data will be categorised and grouped (e.g., by types of intervention facilitators) and where possible, a quantitative description will be provided (e.g. total number of studies reported across reviews).

### Critical appraisal

The risk of bias and quality of methodological results for the included reviews will be evaluated using a standardised appraisal tool specifically designed for the appraisal of systematic reviews and research syntheses [29]. One team member will initially conduct an appraisal which will be cross-checked by a second team member. Any discrepancies will be discussed by team members and resolved.

## Results

To date the search and study selection process is underway, with a search to be re-run in January 2024 to encompass the full search period. A preliminary extraction method has been developed, tested and discussed among the team to help refine the process. It is anticipated that the study will be completed by April 2024.

## Discussion

There is a clear need to ensure that carers have access to evidence-based programs that can effectively support their wellbeing as they care for their loved one. Current models of cancer care rely heavily on the work of carers, and given the growing burden of cancer worldwide [30], this caregiving work is also vital to health system sustainability. This meta-review will facilitate improved understanding of the evidence-base, enabling better identification of research strengths, limitations, and gaps. It will also enhance navigation of the literature, allowing researchers, clinicians, and policymakers to more readily review evidence relevant to them [18, 19], in turn supporting the translation of evidence into practice.

## Supporting information

please see Supplementary file 1

## Data Availability

The manuscript does not contain any data.

https://www.crd.york.ac.uk/prospero/display_record.php?RecordID=403219

## Acknowledgements

Dr Bróna Nic Giolla Easpaig is supported by a Charles Darwin University IAS Rainmaker Start-up Grant. Dr Ursula Sansom-Daly is supported by an Early Career Fellowship from the Cancer Institute NSW (ID: 2020/ECF1163). Dr Rebekah Laidsaar-Powell is supported by an Early Career Fellowship from the Cancer Institute NSW (ID: 2022/ECF1457). Associate Professor Reema Harrison is supported by a Career Development Fellowship from the Cancer Institute NSW (ID: 2021/CDF1104).

## Ethical approval

Ethical approval is not required for this manuscript. The manuscript does not contain any data.

## Author contributions

R.H., B.N., B.N.G.E and J.J. conceptualisation the study. Contributions were made by the full authorship team to provide ongoing guidance about the focus and direction of the study, including in refining data collection. Screening and preliminary data extraction is being undertaken by R.H., B.N., B.N.G.E., R.L-P. and K.S. BNGE produced an initial draft of the manuscript which was reviewed and revised by the full authorship team. The final version of the manuscript is approved by the full authorship team.

