## Supplementary file 2 for "Supporting carers: Study protocol of a meta-review of psychosocial interventions for carers of people with cancer"

### Supplementary file 2 Database(s): Ovid MEDLINE(R) ALL 2013 to February 03, 2023 Search Strategy:

| **#** | **Searches** |
| --- | --- |
| 1 | exp neoplasms/ |
| 2 | (cancer* or oncology* or neoplasm* or carcinoma* or tumo?r* or malignan* or lymphoma* or melanoma* or leuk?emia or sarcoma).tw. |
| 3 | 1 or 2 |
| 4 | (family or families or parent* or mother? or father? or friend? or relative? or spous* or partner? or husband? or wife or wives or son? or daughter? or offIspring? or sibling? or brother? or sister? or informal or friends or significant other*).tw. and (care* or caring).mp. |
| 5 | caregivers/ or caregiver burden/ |
| 6 | (carer* or caregiv* or care giv*).tw. |
| 7 | exp home nursing/ |
| 8 | exp family/ |
| 9 | or/4-8 |
| 10 | ((family or families or parent* or mother? or father? or friend? or relative? or spous* or partner? or husband? or wife or wives or son? or daughter? or offspring? or sibling? or brother? or sister?) adj4 (support* or inform* or train* or educat* or teach* or coach* or instruct* or advis* or advice* or counsel* or therap* or cbt or program* or psycho* or social or pastoral or spiritual or religio* or self help or selfhelp)).mp. [mp=title, book title, abstract, original title, name of substance word, subject heading word, floating sub-heading word, keyword heading word, organism supplementary concept word, protocol supplementary concept word, rare disease supplementary concept word, unique identifier, synonyms] |
| 11 | problem solving.tw. |
| 12 | social support/ or psychosocial.ti,hw. or psychosocial intervention*.mp. or psychosocial support*.mp. |
| 13 | exp social work/ |
| 14 | exp psychotherapy/ |
| 15 | exp counseling/ |
| 16 | education/ |
| 17 | health education/ |
| 18 | teaching/ |
| 19 | exp "religion and psychology"/ |
| 20 | self help groups/ |
| 21 | self care/ |
| 22 | problem solving/ |
| 23 | professional family relations/ |
| 24 | ((psychosocial adj4 intervention*) or (psychosocial adj4 support*) or (support* adj4 intervention*)).ti. |
| 25 | or/10-24 |
| **26** | **3 and 9 and 25** |
| **27** | **limit 26 to (english language and yr="2013 -Current")** |
| 28 | (systematic review or meta-analysis).pt. |
| 29 | meta-analysis/ or systematic review/ or systematic reviews as topic/ or meta-analysis as topic/ or "meta analysis (topic)"/ or "systematic review (topic)"/ or exp technology assessment, biomedical/ or network meta-analysis/ |
| 30 | ((systematic* adj3 (review* or overview*)) or (methodologic* adj3 (review* or overview*))).ti,ab,kf. |
| 31 | ((quantitative adj3 (review* or overview* or synthes*)) or (research adj3 (integrati* or overview*))).ti,ab,kf. |
| 32 | ((integrative adj3 (review* or overview*)) or (collaborative adj3 (review* or overview*)) or (pool* adj3 analy*)).ti,ab,kf. |
| 33 | (data synthes* or data extraction* or data abstraction*).ti,ab,kf. |
| 34 | (handsearch* or hand search*).ti,ab,kf. |
| 35 | (mantel haenszel or peto or der simonian or dersimonian or fixed effect* or latin square*).ti,ab,kf. |
| 36 | (met analy* or metanaly* or technology assessment* or HTA or HTAs or technology overview* or technology appraisal*).ti,ab,kf. |
| 37 | (meta regression* or metaregression*).ti,ab,kf. |
| 38 | (meta-analy* or metaanaly* or systematic review* or biomedical technology assessment* or bio-medical technology assessment*).mp,hw. |
| 39 | (medline or cochrane or pubmed or medlars or embase or cinahl).ti,ab,hw. |
| 40 | (cochrane or (health adj2 technology assessment) or evidence report).jw. |
| 41 | (comparative adj3 (efficacy or effectiveness)).ti,ab,kf. |
| 42 | (outcomes research or relative effectiveness).ti,ab,kf. |
| 43 | ((indirect or indirect treatment or mixed-treatment or bayesian) adj3 comparison*).ti,ab,kf. |
| 44 | (multi* adj3 treatment adj3 comparison*).ti,ab,kf. |
| 45 | (mixed adj3 treatment adj3 (meta-analy* or metaanaly*)).ti,ab,kf. |
| 46 | umbrella review*.ti,ab,kf. |
| 47 | rapid review*.ti,ab,kf. |
| 48 | (multi* adj2 paramet* adj2 evidence adj2 synthesis).ti,ab,kf. |
| 49 | (multiparamet* adj2 evidence adj2 synthesis).ti,ab,kf. |
| 50 | (multi-paramet* adj2 evidence adj2 synthesis).ti,ab,kf. |
| 51 | (synthesis* adj3 evidence).ti,ab,kf. |
| 52 | or/28-51 |
| **53** | **27 and 52** |
| 54 | (developed countries or european union or oecd).tw,hw,sh. |
| 55 | europe/ or andorra/ or austria/ or belgium/ or exp france/ or exp germany/ or exp united kingdom/ or greece/ or ireland/ or exp italy/ or liechtenstein/ or luxembourg/ or monaco/ or netherlands/ or portugal/ or exp "scandinavian and nordic countries"/ or spain/ or switzerland/ or exp australia/ or new zealand/ |
| 56 | north america/ or exp canada/ or exp united states/ |
| 57 | (united kingdom or england or scotland or wales or denmark or finland or iceland or norway or sweden).tw,hw,sh. |
| 58 | (europe* or andorra or austria or belgium or france or germany or greece or ireland or italy or liechtenstein or luxembourg or monaco or netherlands or portugal or spain or switzerland or australia* or new zealand).tw,hw,sh. |
| 59 | or/54-58 |
| **60** | **53 and 59** |
| 61 | (developing countr* or third world or underdeveloped countr* or under developed countr*).mp. |
| 62 | exp africa/ or americas/ or exp caribbean region/ or exp central america/ or latin america/ or mexico/ or exp south america/ |
| 63 | exp europe, eastern/ or exp transcaucasia/ |
| 64 | antarctic regions/ or exp atlantic islands/ or exp indian ocean islands/ or exp pacific islands/ |
| 65 | New Guinea/ or asia/ or exp asia, central/ or asia, southeastern/ or borneo/ or cambodia/ or east timor/ or indonesia/ or laos/ or malaysia/ or mekong valley/ or myanmar/ or philippines/ or thailand/ or vietnam/ or asia, western/ or bangladesh/ or bhutan/ or india/ or middle east/ or afghanistan/ or iran/ or iraq/ or jordan/ or lebanon/ or oman/ or saudi arabia/ or syria/ or turkey/ or yemen/ or nepal/ or pakistan/ or sri lanka/ or far east/ or china/ or tibet/ or exp korea/ or mongolia/ |
| 66 | (Afghanistan or Africa or Albania or Algeria or Angola or Antigua or Argentina or Armenia or Azerbaijan or Bangladesh or Barbados or Barbuda or Belarus or Belize or Bhutan or Bolivia or Bosnia or Botswana or Bulgaria or Burkina Faso or Burundi or Cambodia or Cameroon or Central African Republic or Chad or Chile or china or Colombia or Comoros or Congo or Costa Rica or Croatia or Cuba or Czech* or Congo or Djibouti or Dominica or Dominican or East Timor or Ecuador or Egypt or El Salvador or Equatorial Guinea or Eritrea or Estonia or Ethiopia or Fiji or Gabon or Gambia or Ghana or Grenada or Guatemala or Guinea-Bissau or Guyana or Haiti or Honduras or Hungary or India or Indonesia or Iran or Iraq or Ivory Coast or Jamaica or Jordan or Kazakhstan or Kenya or Kiribati or Kyrgyzstan or Laos or Latvia or Lebanon or Lesotho or Liberia or Libya or Lithuania or Madagascar or Malawi or Malaysia or Maldives or Mali or Marshall Islands or Mauritania or Mauritius or Mexico or Micronesia or Moldova or Mongolia or Montenegro or Morocco or Mozambique or Myanmar or Namibia or Nepal or New Guinea or Nicaragua or Niger or Nigeria or Korea or Oman or Pakistan or Palau or Panama or Papua New Guinea or Paraguay or Benin or China or Peru or Philippines or Poland or Cape Verde or Georgia or Kosovo or Macedonia or Yemen or Romania or Russia or Rwanda or Saint Kitts or Saint Vincent or Saint Lucia or Sao Tome Principe or Saudi Arabia or Senegal or Serbia or Seychelles or Sierra Leone or Slovak* or South Africa or Solomon Islands or Somalia or Sri Lanka or Sri-Lanka or Sudan or Suriname or Swaziland or Syria or Tajikistan or Tanzania or Thailand or Togo or Tonga or Trinidad or Tobago or Tunisia or Turkey or Turkmenistan or Uganda or Ukraine or Uruguay or Uzbekistan or Vanuatu or Venezuela or Vietnam or Samoa or Zambia or Zimbabwe).af. |
| 67 | (low income countries or middle income countries).mp. [mp=title, book title, abstract, original title, name of substance word, subject heading word, floating sub-heading word, keyword heading word, organism supplementary concept word, protocol supplementary concept word, rare disease supplementary concept word, unique identifier, synonyms] |
| 68 | 66 or 67 |
| 69 | 53 not 68 |
| **70** | **60 or 69** |
